# A multi-site, multi-participant magnetoencephalography resting-state dataset to study dementia: The BioFIND dataset

**DOI:** 10.1101/2021.05.19.21257330

**Authors:** Delshad Vaghari, Ricardo Bruna, Laura E. Hughes, David Nesbitt, Roni Tibon, James B. Rowe, Fernando Maestu, Richard N. Henson

## Abstract

Early detection of Alzheimer’s Disease (AD) is vital to reduce the burden of dementia and for developing effective treatments. Neuroimaging can detect early brain changes, such as hippocampal atrophy in Mild Cognitive Impairment (MCI), a prodromal state of AD. However, selecting the most informative imaging features by machine-learning requires many cases. While large publically-available datasets of people with dementia or prodromal disease exist for Magnetic Resonance Imaging (MRI), comparable datasets are missing for Magnetoencephalography (MEG). MEG offers advantages in its millisecond resolution, revealing physiological changes in brain oscillations or connectivity, before structural changes are evident with MRI. We introduce a MEG dataset with 324 individuals: patients with MCI and healthy controls. Their brain activity was recorded while resting with eyes closed, using a 306-channel MEG scanner at one of two sites (Madrid or Cambridge), enabling tests of generalization across sites. A T1-weighted MRI is provided to assist source localisation. The MEG and MRI data can be formatted according to international BIDS standards, and analysed freely on the DPUK platform (https://portal.dementiasplatform.uk/Apply).

## Background & Summary

Alzheimer’s disease (AD) is an age-related neurodegenerative disorder that is characterised by progressive dementia, from mild memory impairment to global cognitive dysfunction and eventually death ^1^. According to the World Alzheimer report in 2019, there are 50 million people in the world with dementia, which is likely to rise to 152 million people by 2050 ^2^. This prevalence accentuates the need for reliable biomarkers that are sensitive to early stages of the disease. Although there is currently no cure for AD, early detection may enable more effective management and the ability to prevent or delay dementia. Biomarkers that are accurate, safe, and sensitive to the specific brain changes in dementia are required to accelerate and increase power for early phase clinical trials.

Here we consider the challenge of Mild Cognitive Impairment (MCI), which is commonly a prodromal state of AD, with a high probability of progression to dementia ^3,4^. MCI is defined by cognitive symptoms and performance on cognitive tests, with or without specific biomarker evidence of underlying AD pathology ^5^. Although a prodromal disorder, patients may have subtle brain changes that are identifiable with neuroimaging. The dominant form of neuroimaging is Magnetic Resonance Imaging (MRI), which is most often used clinically to measure brain structure, particularly the volume of grey-matter in brain regions susceptible to AD, such as in the medial temporal lobes ^6^. However, atrophy is a late pathological stage of neurodegenerative disease, occurring potentially many years after molecular and physiological changes. While functional change can be quantified by functional-MRI, the latter is subject to neurovascular confounds, motion artifacts and low reliability.

Magnetoencephalography (MEG) has been proposed as a valuable alternative tool for functional biomarkers of early stage AD. MEG has better temporal resolution to measure brain function, is reliable across sessions ^7–9^ and is not confounded by neurovascular variance. While Electroencephalography (EEG) can also measure neural activity directly like MEG, it does not offer the same spatial resolution for early, localised effects of disease or changes in functional connectivity (see ^10,11^, for more detailed discussion of the potential advantages of MEG for detecting AD).

MEG offers a large set of potential data features that might differentiate individuals with MCI from healthy controls. These features may be limited to specific frequencies of oscillatory activity, specific brain regions, or the functional connectivity between brain regions. The spatiotemporal complexity of MEG is well suited for machine learning techniques to identify the features that enable classification of MCI ^12^. However, these techniques typically need large number of cases to train and test the classifiers. While large datasets of MRI scans of MCI cases have been made available to the community (e.g, ^13,14^), comparable datasets of MEG are required. Following the recent “BioFIND” project funded by the European Union’s Joint Programming For Neurodegenerative Research initiative ^15^, we combined 168 MEG datasets from a number of ongoing dementia projects at the University of Cambridge, England, and the Centre for Biomedical Technology, in Madrid, Spain. Since then, we have added further data, bringing the total to 324 participants, approximately half of whom had MCI (according to NIA-AA criteria ^16^) while the rest were healthy controls. Most participants had 10 minutes of resting-state MEG (minimum 2 minutes), plus a T1-weighted structural MRI scan (for 309 participants). The MRI can be used to help localise the cortical sources of the MEG data, and also to compare classification based on MEG with that based on the more commonly used structural MRI.

Here we describe this extended BioFIND^17^ dataset of MEG, in the hope that it will allow others to investigate aspects of brain function that differ in MCI patients versus controls, so as to identify potential biomarkers for early AD. We will continue to expand the dataset in future, as new cases are tested at our laboratories, and welcome requests to contribute MEG datasets from other MEG laboratories.

## Methods

### Participants

The 324 participants consist of 158 people with clinically diagnosed MCI and 166 controls, recorded at one of two sites: 1) the MRC Cognition & Brain Sciences Unit (CBU) at the University of Cambridge, and 2) the Laboratory of Cognitive and Computational Neuroscience (UCM-UPM) at the Centre for Biomedical Technology (CTB), Madrid. The participants were pooled over a number of different projects, each approved by local Ethics Committees and following the 1991 Declaration of Helsinki. Participants consented to de-identified data collection and sharing for research purposes.

The 68 MCI patients scanned at Cambridge were recruited from specialist memory clinics at Cambridge University Hospitals NHS Trust; the 91 controls were selected as those with similar age and sex distribution from the population-derived CamCAN cohort of healthy people from the same geographic region ^18^; www.cam-can.org). The 90 patients and 75 controls from Madrid were recruited from the Neurology and Geriatric Departments of the University Hospital San Carlos. The MCI diagnosis was determined with intermediate probability according to the National Institute on Aging–Alzheimer Association criteria ^16^, i.e., given by a clinician based on clinical and cognitive tests, self- and informant-report, and in the absence of full dementia or obvious other causes (e.g., psychiatric). For some patients, there was additional biomarker evidence of atrophy from MRI or long term follow up and genotyping for the *APOE* ε4 allele. Note that the MRI (and MEG) data provided here were research scans following diagnosis, and were not used to inform the diagnosis, though other similar (T1-weighted) clinical MRIs may have been used by the diagnosing clinician. For a subset of MCI patients, we indicate whether or not they subsequently progressed to dementia (probable AD), according to their managing clinician.

The distributions of participant sex, age, education, and score on a cognitive test for dementia - the Mini-Mental State Examination (MMSE) - are shown in Table 1. While MMSE may lack sensitivity to MCI, its widespread use, approval as a clinical trial outcome, and multiple language versions make it a suitable as a screening tool. While t-tests confirmed that the MCI group was slightly older and less well educated on average, there was considerable variance across patients and appreciable overlap between them and the controls, enabling subgroup matching where relevant to future analyses. The MCI group scored lower on the MMSE, with most below the common clinical threshold of 27 ^19.^ The data were acquired about a year earlier for the patients than controls, though again there was overlap in the acquisition year. A Wilcoxon rank sum test showed that the duration of the median MEG recording duration was longer in controls. It is therefore recommended that data are trimmed so that the same duration is used for all participants. While there is no obvious reason why data quality should change over years, we also provide empty-room data for each year and site, to enable estimation of changes in ambient noise levels. There was no significant difference between groups in the time of day of the scan, nor in the mean or standard deviation of their head motion during the scan.

**Table 1.** Participant characteristics

| Data Characteristic | | Groups | | T/$\chi^2$-test |
| --- | --- | --- | --- | --- |
| | | Controls | MCI | T/$\chi^2$ and p value |
| 1 | Site (CBU/CTB) | 91/75 | 68/90 | $\chi^2 = 4.04$ p = 0.04 |
| 2 | Sex (M/F) | 82/84 | 80/78 | $\chi^2 = 0.01$ p = 0.91 |
| 3 | Age (years) | 71.3 (7.0) | 72.9 (6.7) | T = -2.07 p = 0.04 |
| 4 | Education (years) | 14.5 (4.4) | 10.8 (5.3) | T = 6.70 p <.001 |
| 5 | MMSE (/30) | 28.8 (1.2) | 26.1 (2.8) | T = 11.11 p <.001 |
| 6 | Recording Year | 2013.8 (2.4) | 2012.4 (2.5) | T = 5.37 p <.001 |
| 7 | Recording Hour (24h) | 12.8 (2.4) | 12.6 (2.1) | T = 1.13 p = 0.26 |
| 8 | Mean of head translation (mm) | 1.9 (1.8) | 2.3 (2.1) | T = -1.59 p = 0.11 |
| 9 | Standard deviation of head translation (mm) | 1.1 (1.1) | 1.2 (1.1) | T = -1.03 p = 0.30 |
| 10 | Recording Duration (seconds) | 481.5 (262) | 180.0 (305) | Z = 4.19 p <.001 |

*Means have standard deviation in parentheses for items 1 to 9. Medians have interquartile range in parentheses for item 10. Abbreviations: CBU, Cognition & Brain sciences Unit; CTB, Centre for Biomedical Technology; MMSE, Mini-Mental State Examination; MCI, Mild Cognitive Impairment; M, Male; F, Female.*

Note that comparable resting-state MEG data (and T1-weighted MRIs) acquired at the CBU site are also available for approximately 600 healthy participants (aged 18-88 years) via the CamCAN website: https://camcan-archive.mrc-cbu.cam.ac.uk/dataaccess/. These data could be used with machine learning to characterise healthy MEG data, or predict age, which could then be tested on the present patient data.

### Resting-state Protocol

The MEG data were recorded while participants were asked to keep their eyes closed, instructed to think of nothing specific but not fall asleep. The duration of these recordings varied from 2 to 13.35 minutes. Some of the corresponding data-files were extracted from longer raw files, recorded while participants performed other tasks. The time of day and year of the recordings are also provided in the participants.tsv file, but the precise date of the recording was stripped from the raw data file using “mne_anonymze”, in order to further protect participant identity.

### M/EEG data acquisition

MEG recordings were collected continuously at 1 kHz sample rate in magnetically shielded rooms using an Elekta Neuromag Vectorview 306 MEG system (Helsinki, FI). This system includes two orthogonal planar gradiometers and one magnetometer at each of 102 locations around the head. For most participants, bipolar electrodes were used to record the electro-oculograms (EOG), for vertical and/or horizontal eye-movements (though such movements are less common with eyes closed), as well as the electro-cardiogram (“ECG”). When present, these correspond to EEG channels EEG061 (HEOG), EEG062 (VEOG) and EEG063 (ECG); see ‘Data records’ section). For a smaller subset of CBU participants, an additional 70 channels of nose-referenced, unipolar EEG were recorded, but we do not analyse these data here.

To monitor head position throughout the scan, head position indicator (HPI) coils were attached to the scalp and detected by the MEG machine (energized at frequencies above 150 Hz in CTB and above 300Hz in CBU). Prior to the scan, a Fastrak digitizer (Polhemus Inc., Colchester, VA, USA) was used to record locations of the HPI coils, in addition to three anatomical fiducials, for the Nasion, Left and Right Pre-Auricular points (LPA and RPA respectively), plus approximately 100 points across the scalp (to help coregistration with the MRI).

Additional “empty-room” recordings are available for the CBU and CTB without the participant present, which can be used to extract information about the typical environmental magnetic noise. We do not present analyses of these data here.

### MEG pre-processing

In addition to the raw data, we also provide versions that have been de-noised using MaxFilter 2.2.12 (Elekta Neuromag). This entailed: (i) fitting a sphere to the digitized head points, excluding those on the nose, and using the centre of this sphere, together with location of sensors, to define a spherical harmonic basis set for Signal Space Separation (SSS) in order to remove environmental noise (using default number of basis functions), ii) calculation of head position every 1s (though motion was not corrected), and (iii) interpolation of bad channels. The log files produced by MaxFilter are also provided for each participant (see ‘Data Records’ section).

### MRI data acquisition

T1-weighted MRIs for participants tested at the CBU were acquired on either a Siemens 3T TIM TRIO or Prisma using a magnetization-prepared rapid gradient echo (MP-RAGE) pulse sequence. The T1-weighted MRI for participants tested at the CTB were acquired on a General Electric 1.5 Tesla MRI using a high-resolution antenna with a homogenization PURE filter. These images were converted to NIfTI format, and de-faced to protect the participant’s identity.

## Data Records

The data are summarised on the DPUK cohort website (https://portal.dementiasplatform.uk/CohortDirectory/Item?fingerPrintID=BioFIND or https://doi.org/10.48532/007000) and the full set of files can be accessed via the DPUK’s analysis platform: https://portal.dementiasplatform.uk/Apply. There is an application form in order to apply for cohort data access, which once submitted, enables access and analysis through the portal^1^. Within a few days after submission, an instruction email with a necessary credentials will be sent to users by DPUK.

DPUK required the files to be originally uploaded in XNAT format (https://www.xnat.org/). However, we subsequently converted them to the BIDS format using the script called Xnat2Bids.m, which can be found in the GitHub repository that accompanies this paper (https://github.com/delshadv/BioFIND-data-paper/). The Brain Imaging Data Structure (BIDS) format (version 1.4.1; http://bids.neuroimaging.io; see ^20,21^) is an international, community-effort that is more specifically designed for MEG (and MRI) data.

BIDS describes a way of organizing neuroimaging data by defining directory structures, a file naming scheme and file formats. The current version of BIDS does not specify how to handle multi-centre studies, so we simply combined data from both sites into the same directory and identified the site for each participant in the ‘participants.tsv’ file. The present data passed the BIDS validator (https://bids-standard.github.io/bids-validator/).

According to BIDS, data are stored in their native format, and meta-data are stored in “sidecar” text files (.json, .txt, .tsv, etc.) for both human- and computer-readable format.

The top-level directory contains two separate BIDS directory: ‘MCIControls’ (approximately 319GB) and ‘TravelBrains’ (approximately 11GB) (Figure 1a). The ‘MCIControls’ directory includes 324 separate sub-directories (Figure 1b), one per participant, coded ‘sub-Sub’ followed by four digits for the unique participant number, matching the ‘participants.tsv’ file (see below). These directories contain the raw MEG+MRI data; the additional maxfiltered MEG data (see above) are stored in a mirrored format in the ‘derivatives’ sub-directory; furthermore, empty room data are placed within a directory called ‘sub-emptyroom’.

**Figure 1.**
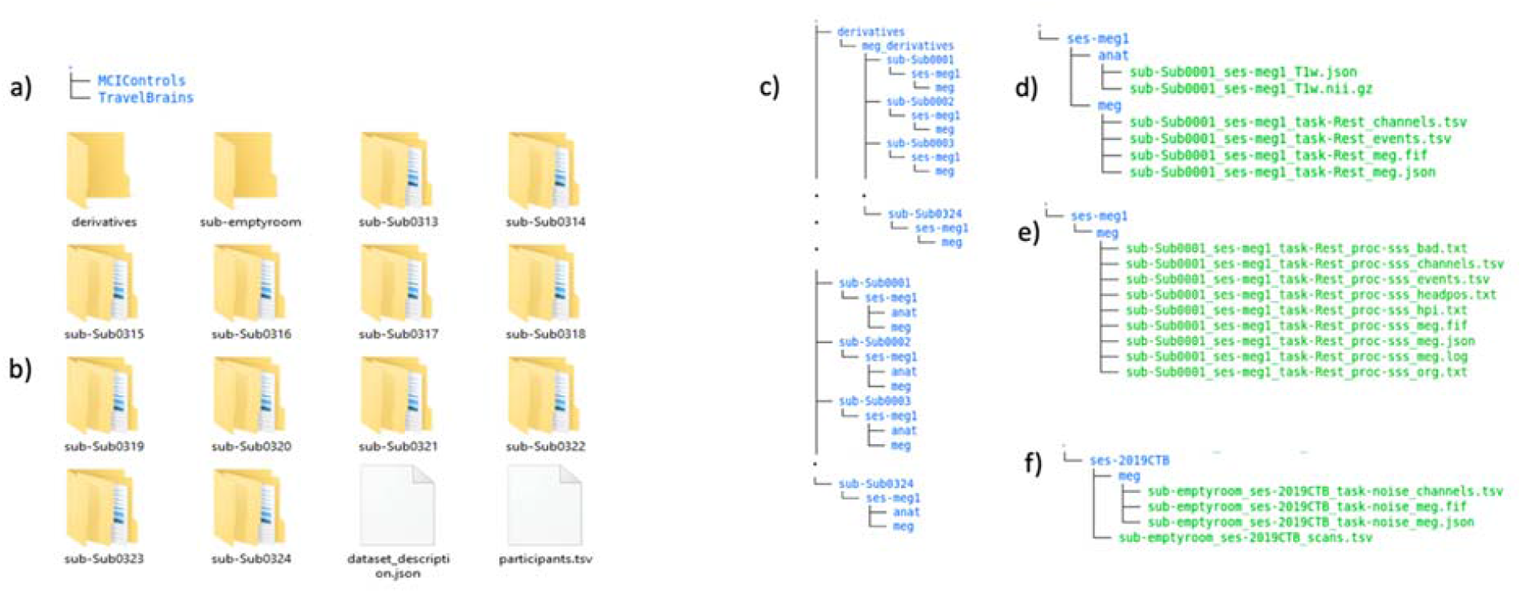
a) Content of Top-level directory b) Organization of MCIControls directory; c) Content of participant-specific directories; d) Content of MRI and MEG data directories for each participant; e) Content of maxfiltered-MEG data directories; f) Content of sub-emptyroom directory

### MCIControls BIDS directory

The MCIControls directory also contains the following files:

The ‘participants.tsv’ file is a tab-separated text file that lists all the participants and associated information, as described in Table 2. Missing data are indicated by ‘n/a’.

**Table 2.**
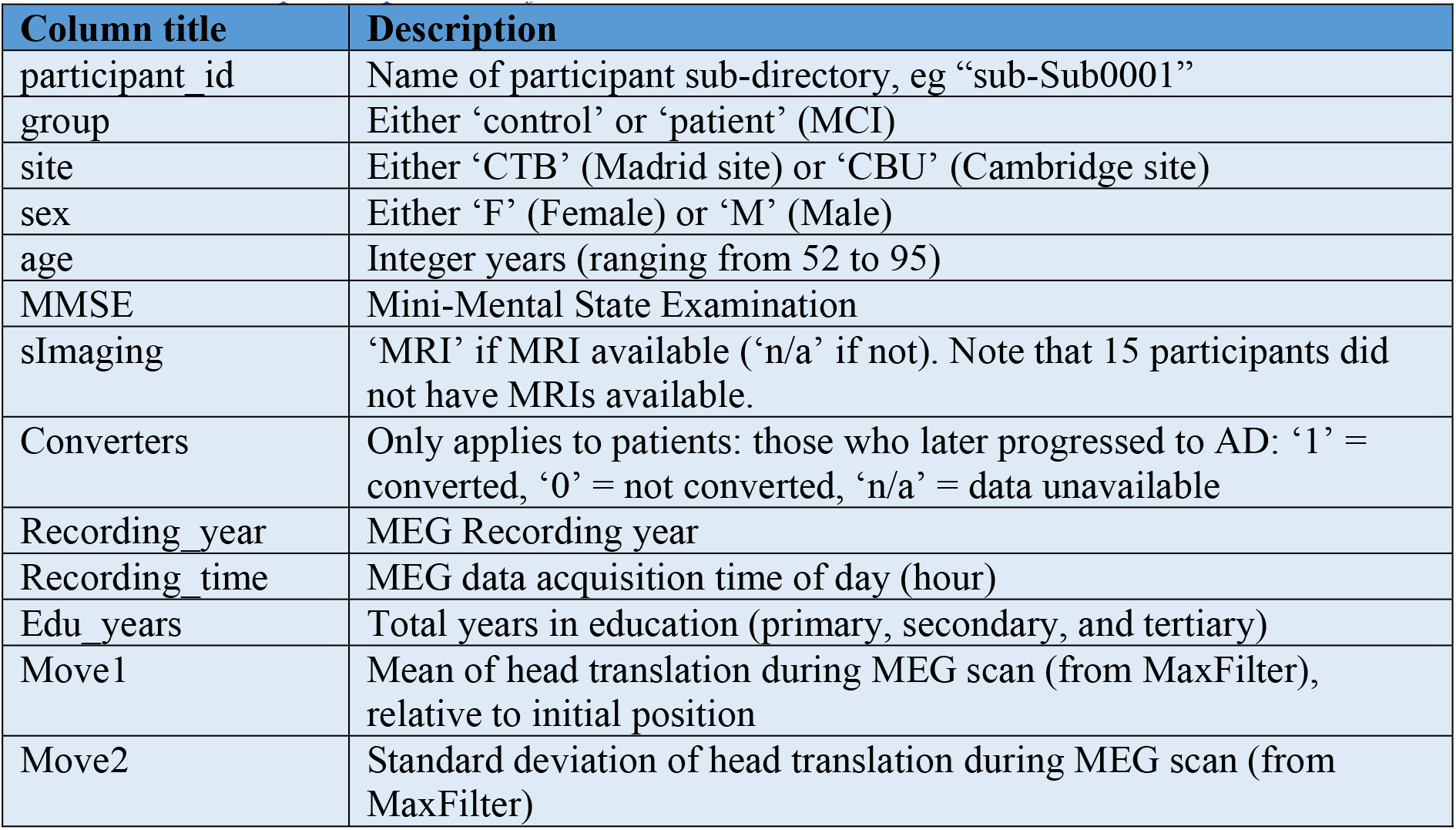
Fields in participants.tsv file

The ‘dataset_description.json’ is a JSON text file describing the dataset including the name, license, authors and how to acknowledge.

### Participant directories

As required by BIDS, within each participant’s ‘sub-Sub####’ directory (where # means one digit) is a sub-directory with a session name and number, in this case always ‘ses-meg1’ (since there is currently only one scanning session per participant). In this session directory are two further sub-directories, ‘anat’ and ‘meg’, which contain the anatomical MRI and MEG data respectively (Figure 1c).

The ‘anat’ folder contains the T1-weighted MRIs, stored in compressed (using GNU zip) NIfTI files, i.e, ‘*.nii.gz’ (where * represents some number of text characters). The international NIfTI format is read by many free software packages. The file name codes the participant number (‘sub-Sub####’), session (‘ses-meg1’) and data type (‘T1w’). Note that the faces on the MRIs imagers were removed using a FreeSurfer ^22^ function (https://surfer.nmr.mgh.harvard.edu/fswiki/mri_deface). There is also a sidecar *.json file, created by hand, which is a text file containing useful meta-data about the T1 image, such as the anatomical MRI coordinate system, and in particular for MEG coregistration, the manually-defined MRI indices for the Nasion, left peri-auricular (LPA) and right peri-auricular (RPA) fiducials. For CBU data the LPA and RPA refer to pre-auricular points; For CTB data the LPA and RPA refer to a point anterior to the tragus. Photographs provided in BIDS directory (Figure 1b).

The ‘meg’ folder contains the raw MEG data, in the native “FIFF” format developed by Neuromag (Elekta Instrumentation AB Stockholm). This format can be read by many free software packages. This file contains data from all the MEG channels, and additional EEG, EOG, ECG and several other miscellaneous channels (of no interest). Note that the precise date and time of recording were scrambled in the FIFF file using the “mne_anonymize” function (https://mne.tools/dev/generated/commands.html#mne-anonymize) of the MNE software ^23^.

Prior to this, the time of day and recording year were extracted and added to the ‘participants.tsv’ file, in case of relevance to the MEG data.

In addition, there are three accompanying sidecar files: *event.tsv, *channel.tsv and *meg.json (Figure 1d). Although resting-state does not have any events, the *event.tsv is included to specify onset and duration of resting state recordings. The *channel.tsv file lists all the channels present in the data, while the *meg.json file encompasses other information about MEG acquisition parameters. These sidecar files were created by the ‘data2bids’ function (http://www.fieldtriptoolbox.org/reference/data2bids/) of the FieldTrip software ^24^.

### Derivatives directory

The BIDS ‘derivatives’ sub-directory contains versions of the data that have been processed in some way. Because MaxFilter ^25^ (https://imaging.mrc-cbu.cam.ac.uk/meg/Maxfilter_V2.2) is proprietary software, we provide versions of the MEG data that have been run through MaxFilter to remove common MEG noise sources (see “MEG pre-processing” section above). The maxfiltered version of each subject’s MEG data is present in a FIFF file (sub-Sub####_ses-meg#_task-Rest_proc-sss_meg.fif) in the corresponding participant directory (Figure 1e). The files have the same name as the original raw MEG data, except for the addition element “proc-sss”, which indicates that the data have been processed with Signal-Space Separation (the method implemented in the MaxFilter software).

In addition to the *proc-sss_meg.json, *proc-sss_channel.tsv and *proc-sss_event.tsv files described above for the raw data, we also provide some additional text files that contain meta-data from the MaxFilter software. These are:

sub-Sub####_ses-meg1_task-Rest_hpi.txt – the 3D locations (in MEG space) of the digitized headpoints

sub-Sub####_ses-meg1_task-Rest_proc-sss_org.txt – coordinate of the centre of a sphere (in MEG space, relative to [0 0 0] as the origin of the helmet) fit to the above headpoints (after excluded points on the nose)

sub-Sub####_ses-meg1_task-Rest_proc-sss_bad.txt – lists MEG channels determined as “bad” for each 10s segment of the data (and subsequently corrected by MaxFilter)

sub-Sub####_ses-meg1_task-Rest_proc-sss_headpos.txt – lists the location of the centre of the head every 1s in quaternions, capturing head motion throughout the scan. The mean and standard deviation of head motion (relative to the initial location) have been extracted and put in the ‘participants.tsv’ file.

sub-Sub####_ses-meg1_task-Rest_proc-sss_meg.log – the full log file output by MaxFilter, containing other information relevant to SSS.

### Empty room directory

Empty room MEG files capture the environmental and system noise, and are located in a directory called ‘sub-emptyroom’ in top level of MCIControls. This directory comprises different sessions for different years and acquisition sites, using the coding ‘ses-YYYYCBU/CTB’ (where YYYY means year of recording) followed by site name, which can be CBU or CTB. In each session’s directory, there is a separate *scans.tsv sidecar file (sub-emptyroom_ses-YYYYCBU/CTB_scans.tsv) containing the date and time of the acquisition expressed in ISO8601 date-time format (YYYY-MM-DDThh:mm:ss); the additional ‘meg’ directory within each session includes corresponding *meg.fif, *channels.tsv and *meg.json files (Figure 1f).

## Travelling Brains

Because of the potential importance of differences between scanners/recording sites, which affects multi-site datasets, we also scanned 7 people on both scanners. These were young, healthy controls (not included in Table 1). Note that MRIs are only available for 4 of these travelling participants, and those MRIs come from different sites. Their MEG data are located in a separate BIDS directory named “Travel Brains”, which includes a participant.tsv file including, sex, age, move1CBU/CTB, move2CBU/CTB, Recording_timeCBU/CTB, Recording yearCBU/CTB. Note that in this case, the ‘site’ refers to the laboratory in which MRI scan was acquired (since the MEG data were acquired in both sites). Because each participant has two FIFF files, one per site, there are now two session directories within each participant directory: ‘ses-megCBU’ and ‘ses-megCTB’. Each session directory holds ‘anat’ directory (when available) and ‘meg’ directory, like for main BIDS repository described above.

BioFIND data are accessible in DPUK data access portal at https://portal.dementiasplatform.uk/AnalyseData/AnalysisEnvironment, following completion of the application process (see above).

## Technical Validation

To check data quality, we applied a simple classifier to some example MEG data features, and examined its ability to distinguish MCI patients from controls, adjusting for age, sex, site, mean and standard deviation of head translation during MEG scan and recording time. We focused on the “alpha” frequency band (8-12Hz) that has previously been associated with ageing and dementia ^26^, and examined: 1) power over the 306 MEG channels, 2) power over 38 cortical brain regions and 3) correlation between the amplitude envelopes of each pair of the 38 ROIs (one measure of functional connectivity ^27^).

Figure 2 illustrates the path of each analyses to train classifier. The full code is available here on GitHub repository https://github.com/delshadv/BioFIND-data-paper/, specifically the Matlab files: feature_extraction_test.m, preproc_beamform_ROI.m and repeated_CV.m.

**Figure 2.**
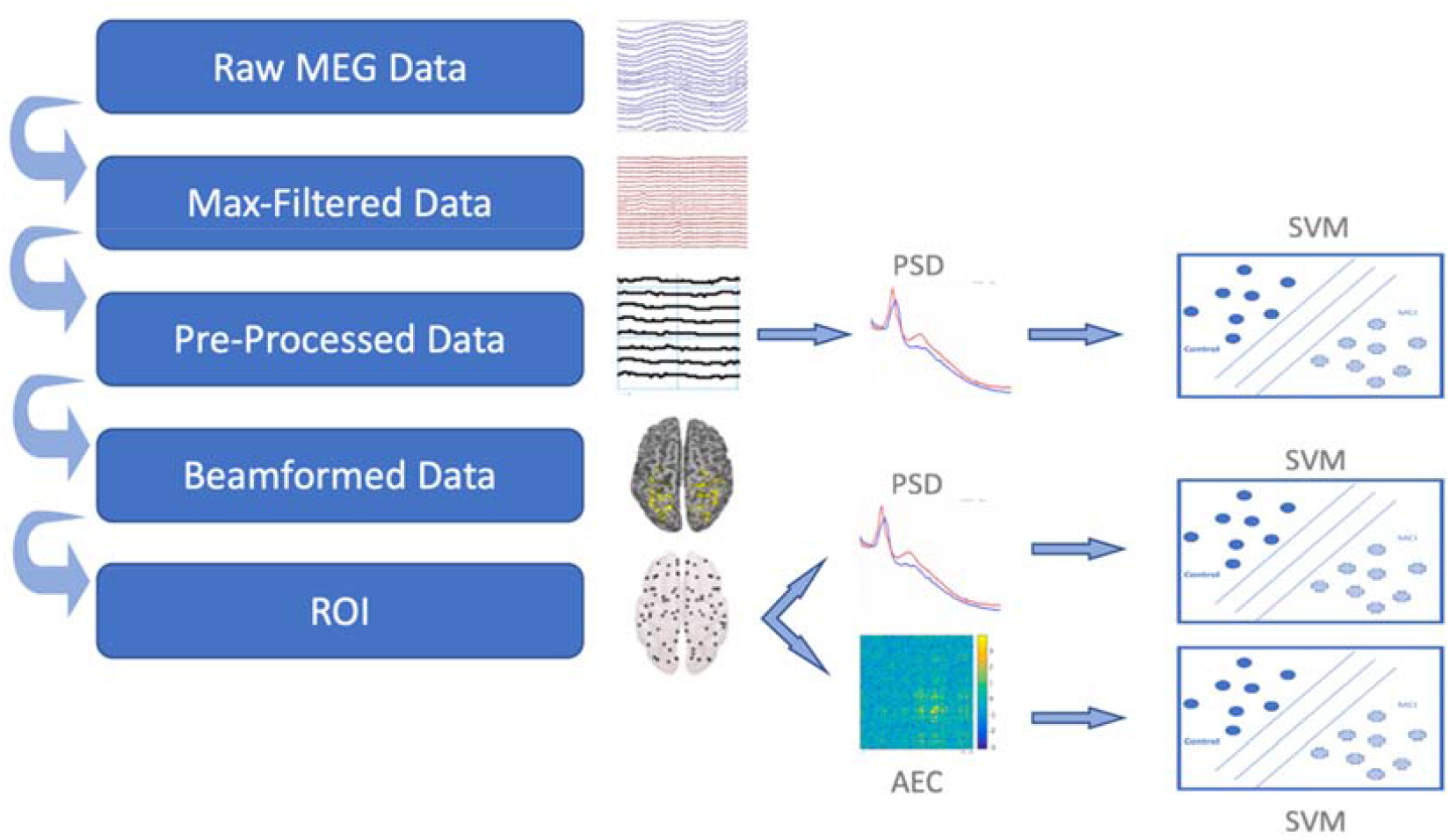
Paths from raw data to classifier trained to distinguish MCI vs Control. Abbreviations: ROI, Region Of Interest; SVM, Support Vector Machine; AEC, Amplitude Envelope Correlation; PSD, Power Spectral Density

The maxfiltered data (in the derivatives directory) were read into MATLAB 2018a (MathWorks, Natick, MA, USA) using the SPM12 software. First, the same length of MEG data for all participants was trimmed to 2 mins to match the lengths of MEG recordings. Data from the 102 magnetometers and 204 gradiometers were then down-sampled to 500 Hz and band-passed filtered from 0.5-150 Hz. The continuous data were then epoched into 2s windows, and epochs that were atypical were marked as bad using the artefact detection function in the OSL toolbox (https://ohba-analysis.github.io/osl-docs/). The remaining epochs were then used to calculate power spectral density (PSD) for each channel every 0.5Hz (using MATLAB’s ‘periodogram’ function) and averaged over epochs. Given the different scaling of the magnetometers and gradiometers, and the fact that the absolute power depends on the position of the cortical sources relative to the sensors (i.e, head position), we calculated relative power by dividing each channel’s PSD by its summed power across frequencies. We then averaged the relative power over frequencies in the range 8-12Hz, to produce 306 features per participant, reflecting the distribution over the scalp of alpha power (relative to total power).

Given that the MEG signal depends on the position and geometry of the head, a more accurate method to estimate brain activity is (in principle) to construct a “forward” model of how the magnetic fields produced by electrical dipoles in the cortex appear at the sensors, based on the shape of the cortex and skull (extracted from an MRI) and information about the position of each sensor relative to the head, and then “invert” that model (with additional assumptions to constrain this ill-posed inverse problem). The first step in this method is to coregister the MRI to the MEG data, which we did by minimising the error between the digitised head-points and the scalp surface extracted from the MRI. We did this using SPM12, after excluding points on the nose since the nose is not always captured in the MRI (the residual error in the 3 anatomically-defined fiducials can be used as an independent measure of coregistration accuracy). Note that this meant excluding the 15 participants for whom no MRI is available.

Once coregistered, we constructed a grid of 3559 cortical sources every 8mm within the brain and estimated how a dipolar source at each location would appear to each sensor using a single-shell forward model. We then estimated those sources using a scalar, Linear-Constrained Miniumum Variance (LCMV) beamformer implemented in the OSL toolbox. Each grid point was then assigned to one of 38 regions of interest (ROIs) based on ^9^, and principal component analysis used to extract a single representative timeseries per ROI. Relative alpha power was then calculated for each ROI, in the same way as for the sensor above, to produce 38 features per participant.

For the final measure of functional connectivity, the resulting 38-node time series were first orthogonalized to avoid spurious correlations using symmetric orthogonalization ^28^. A Hilbert transform from 8-12 Hz was then applied to the ROI timeseries, and the amplitude envelope calculated. This was then downsampled to 1 Hz, and the Pearson correlation coefficient estimated between all pairs of ROIs, producing (38×37)/2=703 unique estimates of functional connections.

The resulting 3 types of MEG data feature are shown in Table 3. These were used to train and test a Support Vector Machine (SVM) ^29^ using cross-validation with 10-folds and performance averaged over 100 permutations. The data features were first adjusted for the covariates of age, sex, site, mean and standard deviation of head translation during MEG scan and recording time by regressing out linear effects of each covariate (missing values of these covariates were imputed using their mean value). Accuracy for the source power measures was approximately 67%, but slightly lower for the sensor power and connectivity measure (possibly due to the greater number of features). Note that one would not expect 100% accuracy, because not all of the MCI group are likely to have AD pathology ^30^. In addition, a proportion of older controls may have latent AD pathology without symptoms. Therefore, this accuracy of 67% provides a benchmark for future work on these data, for example using other types of MEG features (e.g., different frequency bands, different measures of connectivity), other types of machine learning (e.g., random forest, advanced ensemble classifiers, deep learning approaches), and possibly improved methods for pre-processing (de-noising) the MEG data (e.g., using ICA to remove cardiac and other artefacts). Future studies could also distinguish between the subset of MCI patients who subsequently developed probable AD, or explore the role of potential confounds like education.

**Table 3.**
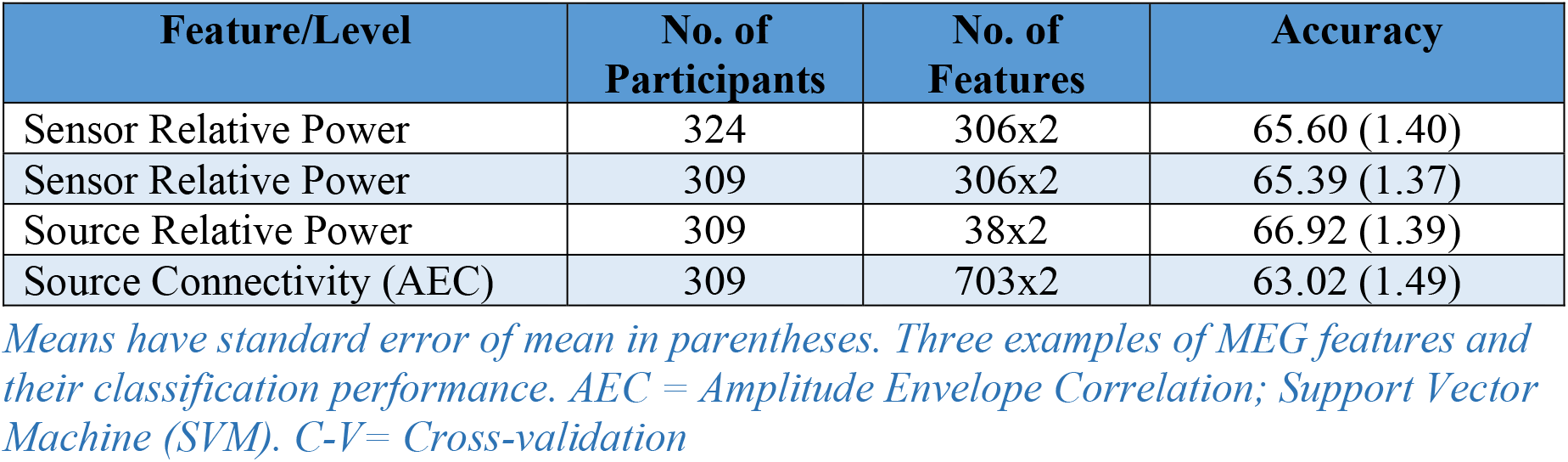
Mean 10-fold cross-validation performance across 100 permutations

## Known exceptions and issues

As noted above, the dataset contains some missing data on some participants, most important of which are: 15 participants missing MRI data; 41 patients without information about subsequent AD progression; 2 participants who only had 2 minutes of data. If such data become available, the dataset will be updated. Moreover, we continue to test new patients and controls in ongoing projects, and when such new data becomes available, we plan to add to the present dataset in future releases.

## Usage Notes

Some of the most common software packages for analysing these data are freely available:

SPM (http://www.fil.ion.ucl.ac.uk/spm/)

OSL (https://ohba-analysis.github.io/osl-docs/)

FieldTrip (http://fieldtrip.fcdonders.nl/)

FreeSurfer (http://surfer.nmr.mgh.harvard.edu/)

MNE (http://martinos.org/mne/)

We request only that researchers acknowledge the authors in any publication arising from these data, and cite this paper and data citation for the source of the data.

## Data Availability

The data are accessible on the UK's Dementia Platform (DPUK): https://portal.dementiasplatform.uk/CohortDirectory/Item?fingerPrintID=BioFIND

https://doi.org/10.48532/007000

## Code Availability

The custom written code to implement all validation analyses is available on GitHub (https://github.com/delshadv/BioFIND-data-paper).

## Acknowledgements

We thank all the participants who contributed their time to these studies, and the scientists who helped collect the data. This work was supported by: EU JNPD (MR/P502017/1), MRC (SUAG/051 G101400; SUAG/046 G101400); the Wellcome Trust (103838) and National Institute for Health Research Cambridge Biomedical Research Centre; two consecutive projects by the Spanish Ministry of Economy and Competitiveness, PSI2009-14415-C03-01 and PSI2012-38375-C03-01. We thank Matthew South, Mark Newbury, Chris Orton, and Emma Squires for help uploading the dataset to DPUK.

## Footnotes

1 For more information, please see application guidance in https://portal.dementiasplatform.uk/Apply.

## Notes

### Competing Interest Statement

The authors have declared no competing interest.

### Funding Statement

This work was supported by: EU JNPD (MR/P502017/1), MRC (SUAG/010-RG91365); the Wellcome Trust (103838) and National Institute for Health Research Cambridge Biomedical Research Centre; two consecutive projects by the Spanish Ministry of Economy and Competitiveness, PSI2009-14415-C03-01 and PSI2012-38375-C03-01.

### Author Declarations

Local Ethics Committees at Cambridge University and Centre for Biomedical Technology (CTB), Madrid.

